# Longitudinal SARS-CoV-2 RNA Wastewater Monitoring Across a Range of Scales Correlates with Total and Regional COVID-19 Burden in a Well-Defined Urban Population

**DOI:** 10.1101/2021.11.19.21266588

**Authors:** Nicole Acosta, María A. Bautista, Barbara J. Waddell, Janine McCalder, Alexander Buchner Beaudet, Lawrence Man, Puja Pradhan, Navid Sedaghat, Chloe Papparis, Andra Bacanu, Jordan Hollman, Alexander Krusina, Danielle Southern, Tyler Williamson, Carmen Li, Srijak Bhatnagar, Sean Murphy, Jianwei Chen, Darina Kuzma, Jon Meddings, Jia Hu, Jason L. Cabaj, John M. Conly, Norma J. Ruecker, Gopal Achari, M. Cathryn Ryan, Kevin Frankowski, Casey R.J. Hubert, Michael D. Parkins

## Abstract

Wastewater-based epidemiology (WBE) is an emerging surveillance tool that has been used to monitor the ongoing COVID-19 pandemic by tracking SARS-CoV-2 RNA shed into wastewater. WBE was performed to monitor the occurrence and spread of SARS-CoV-2 from three wastewater treatment plants (WWTP) and six neighborhoods in the city of Calgary, Canada (population 1.3 million). A total of 222 WWTP and 192 neighborhood samples were collected from June 2020 to May 2021, encompassing the end of the first-wave (June 2020), the second-wave (November end to December, 2020) and the third-wave of the COVID-19 pandemic (mid-April to May, 2021). Flow-weighted 24-hour composite samples were processed to extract RNA that was then analyzed for two SARS-CoV-2-specific regions of the nucleocapsid gene, N1 and N2, using reverse transcription-quantitative polymerase chain reaction (RT-qPCR). Using this approach SARS-CoV-2 RNA was detected in 98.06 % (406/414) of wastewater samples. SARS-CoV-2 RNA abundance was compared to clinically diagnosed COVID-19 cases organized by the three-digit postal code of affected individuals’ primary residences, enabling correlation analysis at neighborhood, WWTP and city-wide scales. Strong correlations were observed between N1 & N2 gene signals in wastewater and new daily cases for WWTPs and neighborhoods. Similarly, when flow rates at Calgary’s three WWTPs were used to normalize observed concentrations of SARS-CoV-2 RNA and combine them into a city-wide signal, this was strongly correlated with regionally diagnosed COVID-19 cases and clinical test percent positivity rate. Linked census data demonstrated disproportionate SARS-CoV-2 in wastewater from areas of the city with lower socioeconomic status and more racialized communities. WBE across a range of urban scales was demonstrated to be an effective mechanism of COVID-19 surveillance.

## 1. Introduction

Since the World Health Organization’s declaration of the coronavirus disease 2019 (COVID-19) a global pandemic, great efforts have been made to monitor for the presence and spread of the Severe Acute Respiratory Syndrome Coronavirus 2 (SARS-CoV-2) around the world. As SARS-CoV-2 RNA is shed in the feces of symptomatic and asymptomatic individuals, its use as a novel investigative tool to understand wastewater-based epidemiology (WBE) of COVID-19 has been aggressively pursued (Amirian, 2020; Cheung et al., 2020; Jiehao et al., 2020; Tang et al., 2020; Wu et al., 2020b; Xagoraraki and O’Brien, 2020; Zhang et al., 2020). Teams from across the world have reported the presence of SARS-CoV-2 RNA in municipal wastewaters (Ahmed et al., 2020; Fitzgerald et al., 2021; Giraud-Billoud et al., 2021; Haramoto et al., 2020; Kumar et al., 2020; La Rosa et al., 2020; Medema et al., 2020; Miyani et al., 2021; Nemudryi et al., 2020; Peccia et al., 2020; Randazzo et al., 2020; Rusiñol et al., 2021; Sherchan et al., 2021; Westhaus et al., 2021; Wu et al., 2021; Wu et al., 2020a; Wurtzer et al., 2020). These studies have shown that WBE is suitable for spatial and temporal monitoring of SARS-CoV-2 across diverse communities (Kitajima et al., 2020), serves as a leading indicator correlating strongly with future diagnosed cases, hospitalizations and deaths (Miyani et al., 2021; Zhu et al., 2021), predicts the burden of undiagnosed infections at the population level (Bivins et al., 2020; Daughton, 2020; Nemudryi et al., 2020; Wannigama et al., 2021) and can play a key role in optimizing public health decision-making (Daughton, 2020).

To date, COVID-19 WBE studies have generally focused on samples collected from wastewater treatment plant (WWTP) influent where these data assess each city as a single, homogenous entity. However, significant regional and social disparities exist within communities and influence COVID-19 incidence risk and associated morbidity/mortality (Karmakar et al., 2021; Mena et al., 2021). A nested monitoring strategy that samples wastewater at different spatial scales within a city (e.g., from municipal zones, down to individual neighborhoods and even buildings) has the potential to provide greater granularity in understanding disease transmission and its associated impact. This study sought to understand how SARS-CoV-2 wastewater measurements collected at a range of scales (extending from neighborhoods – through to an entire city) across an urban metropolis with high levels of public health data integration correlated with clinically diagnosed cases.

Among Canada’s cities, Calgary is the fourth largest and third most ethnically diverse (Calgary, 2020). Calgary is an ideal place to perform WBE modelling of COVID-19 for a number of reasons. (i) Unique relative to other cities, Calgary has separate stormwater and wastewater collection systems, preventing epidemiological signals in wastewater from being diluted by stormwater; (ii) a single municipal utility provider is responsible for waterworks, enabling consistency and greater ease of sampling; (iii) all healthcare and COVID-19 diagnostic testing are performed and data captured by a single publicly funded health-service provider, Alberta Health Services; and, (iv) longstanding relationships between the University of Calgary and the City of Calgary through the Urban Alliance (University-Calgary, 2021) enabled the rapid development and deployment of WBE. These advantages have enabled this transdisciplinary nested approach to longitudinal monitoring for SARS-CoV-2 in wastewater from June 2020 to May 2021 that assessed SARS-CoV-2 RNA signal accuracy at the level of individual neighborhoods, each of Calgary’s three WWTPs, and a flow-adjusted aggregate signal representing the entire city of Calgary.

## 2. Methods and Materials

### 2.1. Wastewater sampling and pre-processing

Calgary has three-wastewater treatment plants WWTPs (Figure 1) that serve an estimated 1,441,268 people in Calgary and surrounding communities (Alberta-Government, 2020). Composite raw wastewater from each of the three WWTPs was collected twice weekly from June 29, 2020, to May 31, 2021 (Table 1). Six neighborhood specific sampling sites (Table 1, Figure 1) were sequentially added to this bi-weekly sample collection schedule to enable more granular geographic assessment of the spatial and temporal variation in SARS-CoV-2 RNA levels throughout the city. Neighborhoods were chosen based on ease of access, size of population, and to represent a diverse cross-section of Calgary.

**Figure 1.**
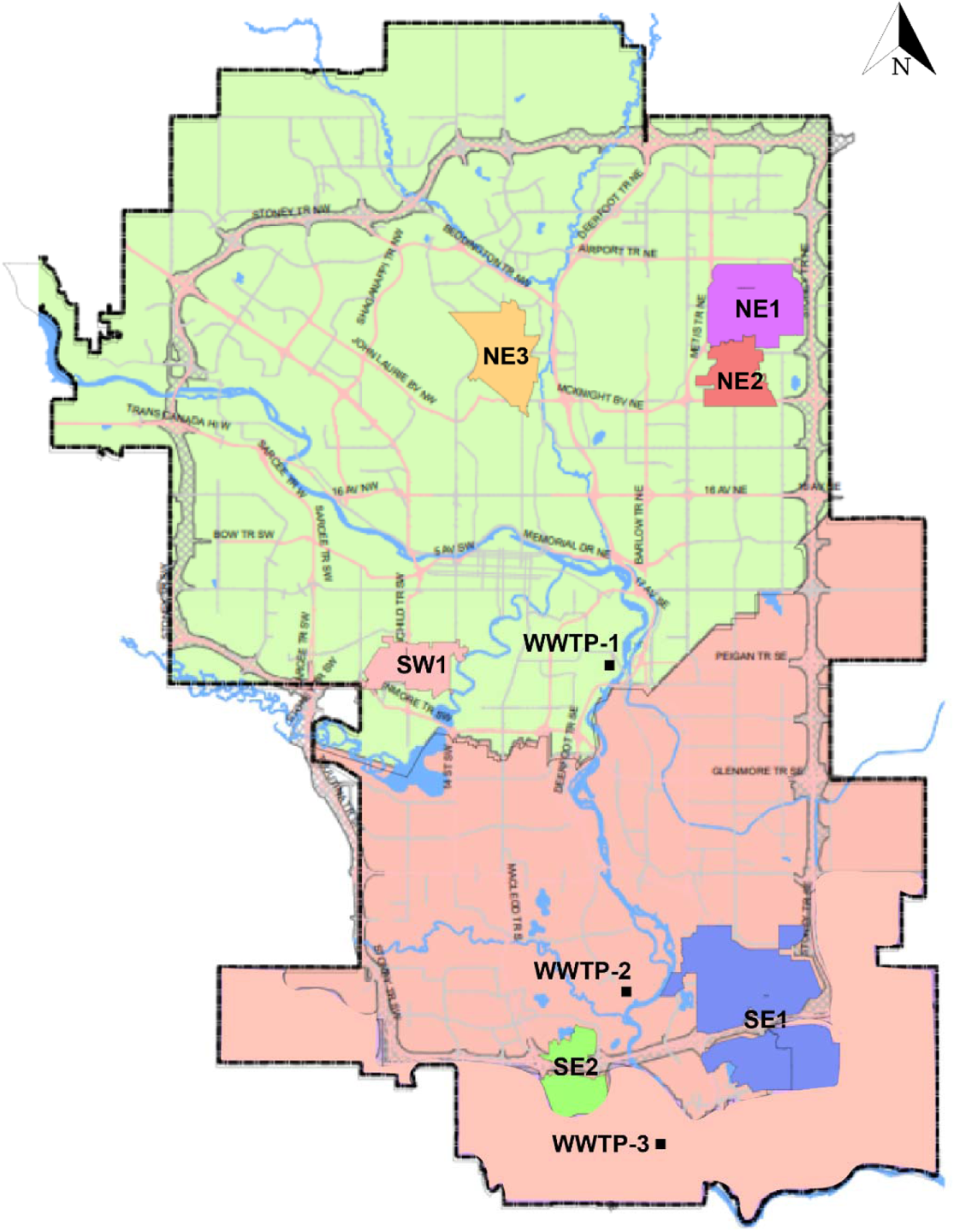
Map of Calgary showing the catchment of the three WWTPs and the six targeted neighborhoods included in the study. Catchment area for the three WWTPs (Table 1) are shaded on the map as follows: WWTP-1 is green, WWTP-2 and WWTP-3 are in orange owing to their shared catchment area. Six neighborhoods are shown by the smaller shaded regions, and are designated by quadrant: NE1, NE2, NE3, SE1, SE2 and SW1.

**Table 1.**
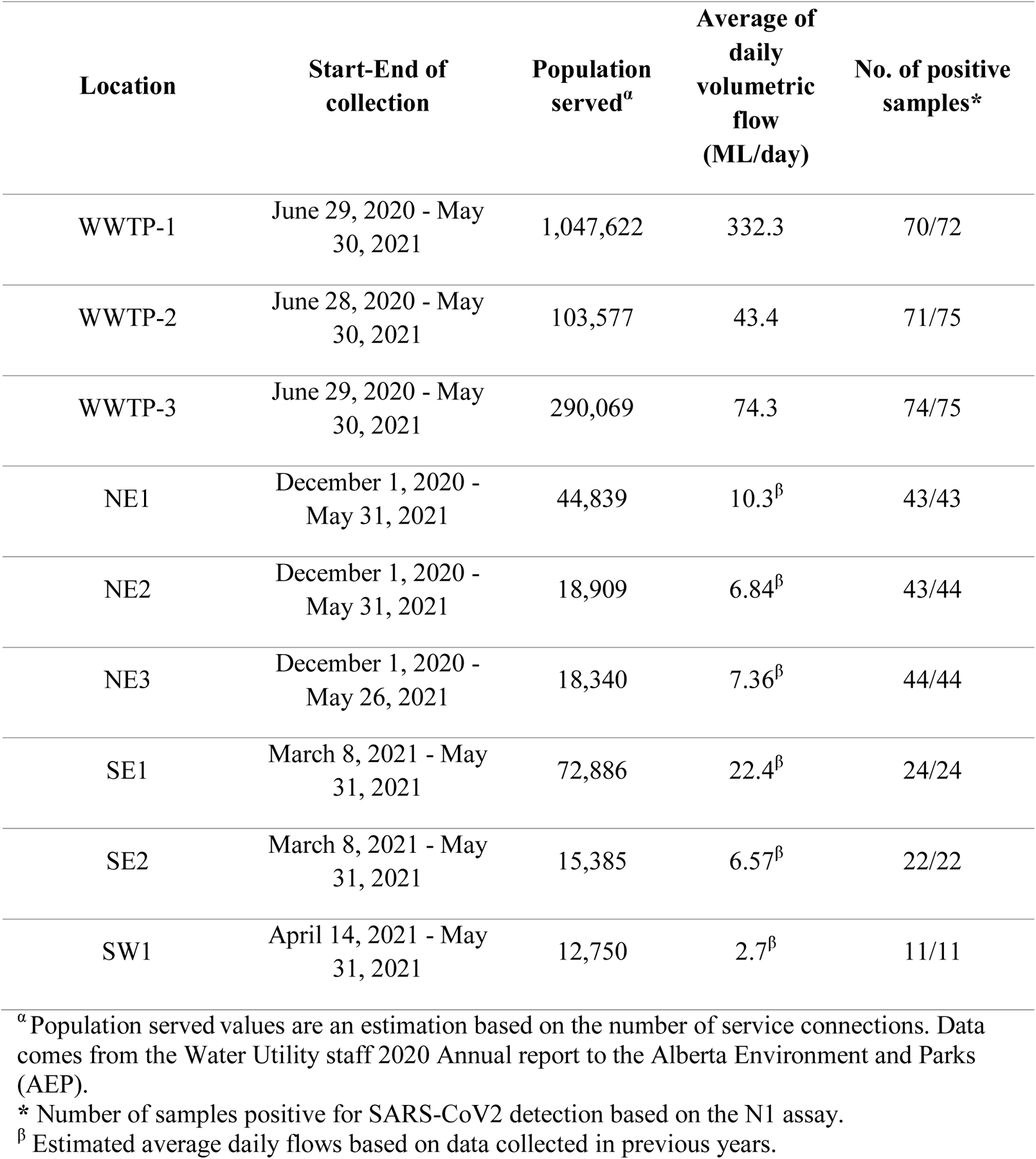
Wastewater treatment plants and neighborhoods monitored for SARS-CoV-2 RNA.

| Location | Start-End of collection | Population served<sup>a</sup> | Average of daily volumetric flow (ML/day) | No. of positive samples* |
| --- | --- | --- | --- | --- |
| WWTP-1 | June 29, 2020 - May 30, 2021 | 1,047,622 | 332.3 | 70/72 |
| WWTP-2 | June 28, 2020 - May 30, 2021 | 103,577 | 43.4 | 71/75 |
| WWTP-3 | June 29, 2020 - May 30, 2021 | 290,069 | 74.3 | 74/75 |
| NE1 | December 1, 2020 - May 31, 2021 | 44,839 | 10.3<sup>β</sup> | 43/43 |
| NE2 | December 1, 2020 - May 31, 2021 | 18,909 | 6.84<sup>β</sup> | 43/44 |
| NE3 | December 1, 2020 - May 26, 2021 | 18,340 | 7.36<sup>β</sup> | 44/44 |
| SE1 | March 8, 2021 - May 31, 2021 | 72,886 | 22.4<sup>β</sup> | 24/24 |
| SE2 | March 8, 2021 - May 31, 2021 | 15,385 | 6.57<sup>β</sup> | 22/22 |
| SW1 | April 14, 2021 - May 31, 2021 | 12,750 | 2.7<sup>β</sup> | 11/11 |

<sup>a</sup> Population served values are an estimation based on the number of service connections. Data comes from the Water Utility staff 2020 Annual report to the Alberta Environment and Parks (AEP).
\* Number of samples positive for SARS-CoV2 detection based on the N1 assay.
<sup>β</sup> Estimated average daily flows based on data collected in previous years.

WWTP and neighborhood wastewater samples were collected by City of Calgary Water Utility Services staff and/or the university research team using a consistent standard protocol. ISCO 5800 or ISCO 6712 portable autosamplers were programmed to collect and store 24-hour composite samples. Sample collection from WWTP-2, WWTP-3 and neighborhood locations were time-weighted, whereas for samples collected from the larger WWTP-1 were flow-weighted. Detailed description of sample collection parameters is described in the Supplemental Material. Immediately after collection, samples were transported to the University of Calgary’s Advancing Canadian Wastewater Assets (ACWA) lab for pre-processing and RNA isolation using a modified version of the Sewage, Salt, Silica, and SARS-CoV-2 (4S) protocol (Whitney et al. 2021). This method enables unbiased recovery of RNA from intact and lysed SARS-CoV-2 as well as from other biological particles (Acosta et al., 2021; Whitney et al., 2021). Samples were thoroughly mixed, subsampled and spiked with 200 µl of a Bovine coronavirus (BCoV) surrogate control. An extraction blank (i.e., UltraPure™ DNase/RNase-Free Distilled Water (Invitrogen)) was included in each batch of processed samples. Extracted nucleic acids were transported on dry ice to the Health Sciences Centre, University of Calgary, for subsequent RT-qPCR analysis.

### 2.2. RT-qPCR analysis

RT-qPCR assays for the SARS-CoV-2 nucleocapsid gene (N1 and N2) and BCoV M gene were performed using a QuantStudio-5 Real-Time PCR System (Applied Biosystems) (Acosta et al., 2021). All RT-qPCR assays were performed in triplicate according to the MIQE guidelines (Bustin et al., 2009). Samples were considered positive for N1 or N2 gene targets if the threshold quantification cycle (Cq) was lower than 40 cycles (Acosta et al., 2021; Medema et al., 2020; Randazzo et al., 2020; Wu et al., 2020a). Quality assurance and quality controls are described in Supplementary Material.

SARS-CoV-2 concentrations were assessed for each individual WWTP and as a composite wastewater volume-weighted city-wide SARS-CoV-2 mass flux. In the latter case, the total mass flux of SARS-CoV-2 was calculated as the sum of the mass flux from each of the three WWTPs, such that the daily SARS-CoV-2 mass flux at each WWTP can be determined as the product of the SARS-CoV-2 concentration and the daily volumetric flow (Table 1).

### 2.3. Demographic and clinical data collection

The population monitored from each sewershed catchment (i.e., those corresponding to individual WWTPs and neighborhoods) were estimated based on Calgary’s 2019 census information (Calgary, 2020). For each day, total-active cases (i.e., confirmed by validated clinical RT-qPCR assays) and newly diagnosed cases were collected by Alberta Health Services (AHS) (https://covid-tracker.chi-csm.ca/) and binned by individual postal codes (using the first three of six digits) as indicated by the home address of newly diagnosed cases. Postal codes corresponding to individual neighborhoods and WWTPs were used to correlate wastewater data with clinical information. In instances where the partial postal code did not overlap entirely with individual sewershed boundaries, the proportion of cases ascribed to the community of interest was based on the proportion of residences served by the sewershed’s geographic area.

Five-day rolling averages of new clinical case numbers were used for correlation analyses. Details on the introduction and relaxation of restrictive public health ordinances instituted by public health officials to control the pandemic were collected from Alberta Health (Alberta-Government, 2020). Socioeconomic status (SES) and ethno-demographic indicators for communities corresponding to sampled sewershed catchment zones were obtained from 2016 Canadian Census (Pinault et al., 2020; Statistics-Canada, 2016), as described in the Supplementary Material. Wastewater monitoring data from this study has been presented publicly in real time by the University of Calgary’s Centre for Health Informatics COVID Tracker (https://covid-tracker.chi-csm.ca/). This study received institutional ethics approval from the Conjoint Regional Health Ethics Board of the University of Calgary (REB20-1544).

### 2.4. Statistical analysis

For each sample and target gene combination, triplicate RT-qPCR was performed, with average values for Cq and quantification (copies/ml) used for statistical analysis. Copies per reaction were converted to copies per unit volume of wastewater using dimensional analysis (Acosta et al., 2021). Pearson’s correlation analyses were conducted for assessing relationships between SARS-CoV-2 WW RNA concentration and public health data for COVID-19 (new cases and total cases). SARS-CoV-2 RNA signals were assessed in aggregate and in two-month blocks owing to profound changes in disease burden and circumstances. These blocks were June to July 2020, August to September 2020, October to November 2020, December 2020 to January 2021, February to March 2021, and April to May 2021. Kruskal-Wallis tests were conducted to determine whether differences in the overall SARS-CoV-2 RNA signal existed between composite data for both WWTP and neighborhood during different time periods. Dunn’s Multiple Comparison Test was used to determine which specific sites differed from others across time. Comparison of Calgary’s wastewater average flow (megaliters (ML) per day) during the sampling period across all WWTPs were conducted using one-way ANOVA. Differences in proportions among categorical data were assessed using Fisher’s exact testing for pairwise comparisons (2-sided). All statistical analyses were conducted in GraphPad’s Prism-8 software (La Jolla, CA).

## 3. Results

### 3.1. WBE at WWTP scale

Wastewater samples were analyzed twice-weekly starting on June 29, 2020 after the peak of Calgary’s “first-wave” of COVID-19, through to May 31, 2021. A total of 223 wastewater samples were collected at the WWTP facilities (73, 75, and 75 for WWTP-1, WWTP-2, and WWTP-3, respectively) (Table 1) (Figure 1). Average WWTP flow during the sampling period varied from 43 to 332 ML/day with significantly different daily median volumetric flow between the facilities (WWTP-1: 323.7 (IQR: 316.1-330.6), WWTP-2: 31.6 (IQR: 29.1-54.6), and WWTP-3: 84.8 (IQR: 70.5-87.7), P<0.001). Only one WWTP sample (0.45%) was excluded during this analysis period owing to low signal in the internal controls (Supplemental Material, Table 3S). Cq for N1 and N2 detection were positively correlated with each other across all WWTPs (Pearson’s r=0.837, CI: 0.789-0.874, P<0.001). The N1 assay was more sensitive than the N2 assay for detecting SARS-CoV-2 RNA in wastewater samples, as reported previously (Acosta et al., 2021). Of the 222 WWTP samples tested using the N1 and N2 assays, 195 samples were positive for both and 215 were positive for N1 (no samples were positive for N2 without also being positive for N1). Compared to N1, N2 had a sensitivity of 90.7%. Accordingly, the N1 assay was considered the standard for this study for all subsequent analyses. The proportion of wastewater samples testing positive for N1 at the WWTP-1, WWTP-2, and WWTP-3 were 97.2%, 94.6% and 98.6%, respectively. RT-qPCR performance is described in Supplementary Material.

In general, SARS-CoV-2 RNA concentrations in all WWTP samples collected between late-June 2020 and mid-October 2020 were low but above the limit of detection. When community cases were less than 2.1 per 100,000 residents, wastewater results were likely to be negative (Fisher’s exact test, P=0.001) (Figure 2). Beginning in mid-October there was an increase in SARS-CoV-2 RNA at all three WWTPs, peaking 12 days ahead of the peak case rate in early December 2020 during Calgary’s second wave of COVID-19. For all time periods the SARS-CoV-2 RNA concentration was highest in WWTP-1 compared to the other two sites (Figure 2A), consistent with postal code data of clinical diagnoses. SARS-CoV-2 RNA concentration between October to November 2020 was 2-4 times higher for WWTP-1 relative to either WWTP-2 (Figure 2B) or WWTP-3 (Figure 2C) during that period, but these differences were not significant (Kruskal-Wallis test, P= 0.110). Differences in SARS-CoV-2 RNA levels between WWTPs were more marked during the December 2020-January 2021 sampling period (Kruskal-Wallis test, P= 0.015) and during the April 2021-May 2021 sampling period (Kruskal-Wallis test, P= 0.002) (Figure 3A), coinciding with Calgary’s second and third waves of COVID-19. To determine how WWTPs differed, Dunn’s multiple comparisons test was performed for these two sampling

**Figure 2.**
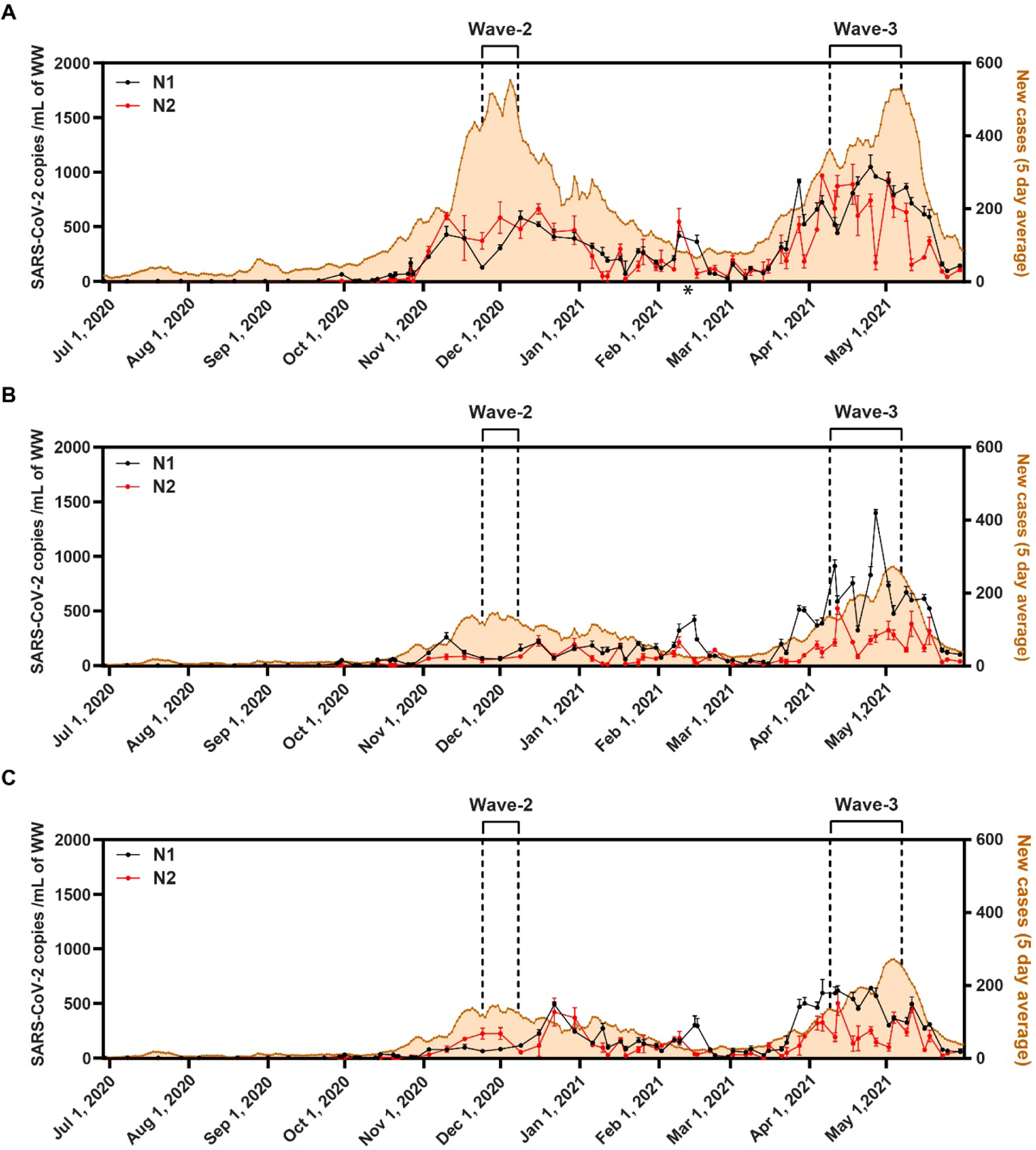
Time series of SARS-CoV-2 RNA concentration for three different wastewater treatment plants. SARS-CoV-2 RNA signal plotted together with daily number of COVID-19 new cases (rolling 5-day average) categorized by three-digit postal codes of diagnosed individuals’ home addresses (orange line). Data were collected from June 29, 2020 to May 31, 2021 from all three WWTPs. SARS-CoV-2 RNA signals are shown for both N1 (black line) and N2 (red line) assays performed on wastewater samples collected twice weekly from Calgary’s three WWTPs (Table 1). Error bars correspond to the standard deviations of the three technical replicates. **A.** WWTP-1, **B.** WWTP-2 and **C.** WWTP-3. Calgary’s second and third waves of COVID-19 are represented as the vertical dotted lines. Asterisks (*) denote the single sample that was not included in the analysis (i.e., WWTP-1 (15/02/2021) due to the presence of inhibitors.

**Figure 3.**
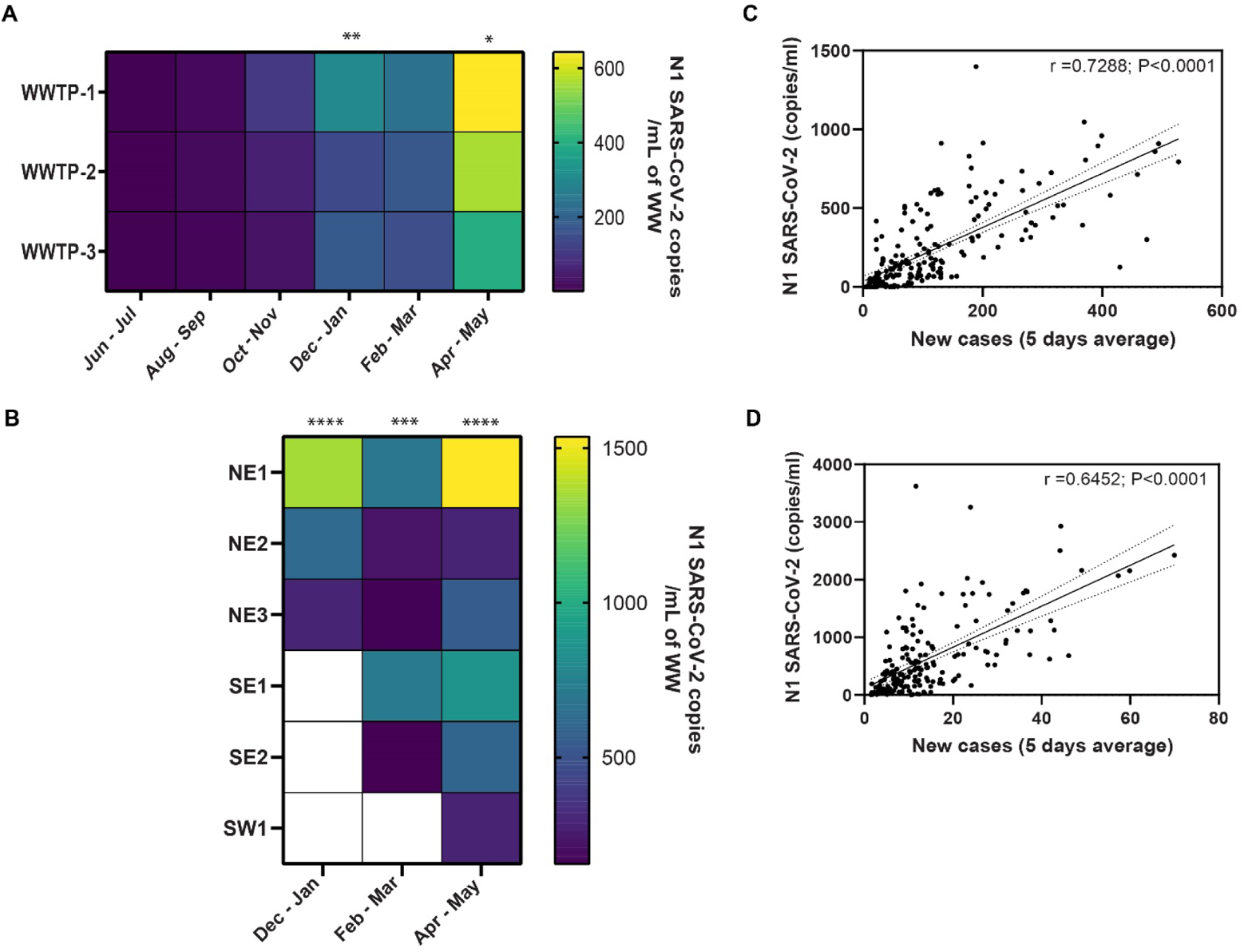
Comparison of SARS-CoV-2 N1 RNA signal at nested geographic scales. **(A)** Comparison of SARS-CoV-2 N1 RNA signals from different WWTPs, and (**B**) neighborhoods. Data were binned into two-month blocks by sampling date (July and August; September and October, etc). White squares denote no sample collection. Asterisks denote values being significantly different among sites when Kruskal-Wallis test was tested (One asterisk (*) denotes P<0.05; two asterisks (**) denote P<0.01; three asterisks (***) denote P<0.001 and four asterisks (****) denote P<0.0001). Correlations between data pairs of new COVID-19 cases organized by three-digit postal codes (5-day average) and SARS-CoV-2 N1 copies/ml in all WWTPs (**C**) or in all neighborhoods sites (**D**). Pearson correlations and 95% prediction intervals (dashed line) on the linear regressions (solid line) are shown in each figure. WWTP: wastewater treatment plant.

periods. During the second wave the concentration of SARS-CoV-2 copies in WWTP-1 was significantly higher than WWTP-2 and WWTP-3 (adjusted p values = 0.003 and 0.019, respectively), and during the third wave WWTP-1 was greater than WWTP-3 (adjusted p value = 0.012).

To determine the relationship between clinically diagnosed COVID-19 and SARS-CoV-2 RNA in wastewater, Pearson correlations were performed using the 5-day average of new cases linked to the three-digit postal codes for the sewershed catchments served by each WWTP. We observed a strong correlation between wastewater N1 signals and new daily cases for all WWTPs when analyzed individually (Table 2). N2 signals also strongly correlated with the number of incident cases in each area (Supplemental Material, Table 4S).

**Table 2.** Correlations between daily COVID-19 cases and S ARS-CoV-2 gene copies/ml in different sampling locations using the N1 assay.

| Location | SARS-CoV-2 gene copies/ml vs new cases (rolling 5-day average)* |  |  |
| --- | --- | --- | --- |
|  | r | P value | 95 % CI |
| WWTP-1 | 0.820 | <0.0001 | 0.726 to 0.884 |
| WWTP-2 | 0.735 | <0.0001 | 0.610 to 0.825 |
| WWTP-3 | 0.707 | <0.0001 | 0.572 to 0.805 |
| NE1 | 0.627 | <0.0001 | 0.402 to 0.780 |
| NE2 | 0.330 | 0.029 | 0.036 to 0.571 |
| NE3 | 0.595 | <0.0001 | 0.362 to 0.758 |
| SE1 | 0.354 | 0.089 | -0.057 to 0.662 |
| SE2 | 0.4285 | 0.046 | 0.0084 to 0.72 |
| SW1 | 0.425 | 0.19 | -0.235 to 0.817 |

\* New cases defined to the three-digit postal code level.

### 3.2. WBE at city-wide scale

To develop a single city-wide mass flux reporting metric, the flux of SARS-CoV-2 RNA for each WWTP was calculated by multiplying the copies/ml by the daily influent flow rate data, and then combining the three WWTPs. City-wide mass flux data was calculated from 42 weeks of observation extending from the end of the first wave through to end of the third wave (Figure 4A). At this city-wide scale, a positive N1 signal was reliably detected during periods where daily new clinical cases exceeded 1.14 per 100,000 people (Figure 4A). The type and timing of implemented public health ordinances are described in the Supplementary Material.

**Figure 4.**
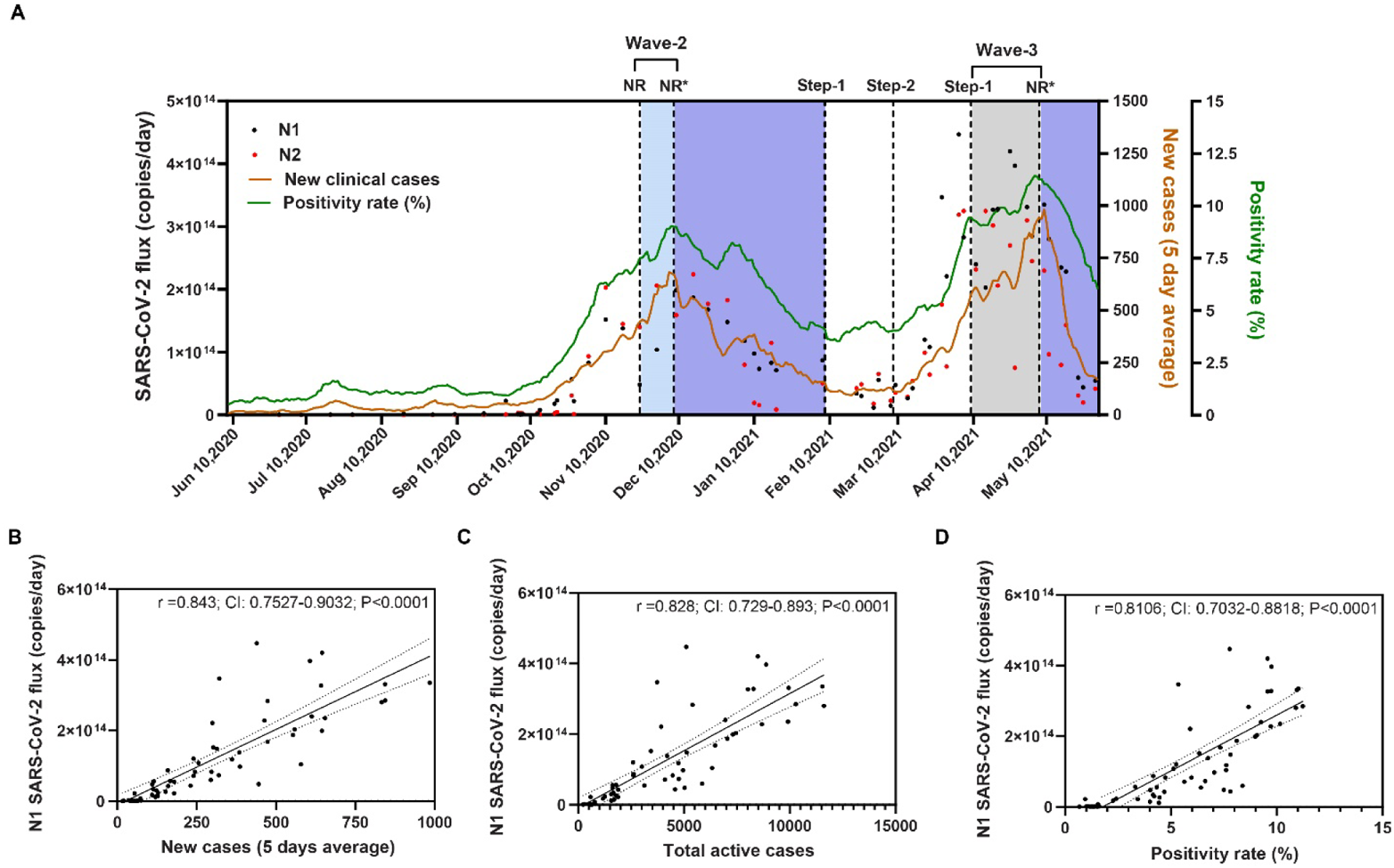
Total Calgary mass flux of SARS-CoV-2 RNA compared to COVID-19 clinically diagnosed cases. **(A)** Daily dynamics of the mass flux of SARS-CoV-2 RNA signal in entire city of Calgary from June 29, 2020 to May 31, 2021 using the N1 (black dots) and N2 (red dots) assays. COVID-19 new cases (5-day average) and clinical test percent positivity rate are represented as orange and green lines. Calgary’s second and third waves of COVID-19 are represented as the blue and gray shaded areas. Vertical dotted lines mark the introduction of new restrictions (NR) on November 24 and December 8, 2020 (stricter restrictions). Some of the restrictions that were implemented during the second wave were partially lifted on February 8, 2021. Further relaxation took place on March 8, 2021. On May 7, 2021, new stricter restrictions (NR*) were once again put in place. Decreases in both the SARS-CoV-2 wastewater signal and clinically diagnosed cases are represented as the purple-shaded areas. Correlations between flow corrected data of the SARS-CoV-2 N1 (copies/day) and new cases (5 days average) (**B**), COVID-19 total active (**C**) and the clinical test percent positivity rate (**D**). Pearson correlations and 95% prediction intervals (dashed line) on the linear regressions (solid line) are shown in each figure. WWTP: wastewater treatment plant, NR: new restrictions, NR*: stricter new restrictions.

We assessed the correlation between SARS-CoV-2 flow corrected signal in wastewater with clinical data (i.e., total-active cases, new daily cases, and test positivity rate) to better understand the natural history of WBE. For the N1 wastewater-signal assay, we observed the strongest correlations between SARS-CoV-2 RNA and clinical data to occur with new daily diagnosed cases (Figure 4B), followed by total active cases (Figure 4C) and test positivity rates (Figure 4D). Similar correlations were observed using N2 assays for the new daily cases (5 day-average) (Pearson’s r=0.7887, CI: 0.6712-0.8676, P<0.001), total active cases (Pearson’s r=0.725, CI: 0.5804-0.8253, P<0.001), and COVID-19 test positivity rate (Pearson’s r=0.7616, CI: 0.632-0.8497, P<0.001).

### 3.3. WBE at neighborhood scale

To test whether wastewater could be used for more granular SARS-CoV-2 surveillance upstream of the WWTPs, we collected samples from different locations within Calgary. Composite 24-h wastewater samples from six separate neighborhoods (Figure 1) were brought online sequentially over 25 weeks, resulting in a total of 191 samples being collected between December 1, 2020, and May 31, 2021 (Table 1, Figure 5). Cumulatively, these six sampling locations represented WW from 183,109 residents (14.2% of Calgary’s population), with the six populations sizes ranging from 12,000 to 72,000 residents (Table 1). Sites were chosen to represent the geographic diversity of Calgary and are notable for significant heterogeneity with respect to Statistics Canada’s indicators of ethno-demographics and socioeconomic status (Figure 6). Three samples (1.6%) were excluded owing to low signal in our internal controls (Supplemental Material, Table 3S). As was observed for WWTPs, the N1 assay was more sensitive than the N2 assay, with 180 samples positive for both targets and 187 positive only for N1 (99.4%). Again, no samples were positive for N2 and not N1. Cq values for N1 and N2 were positively correlated for all neighborhood samples (Pearson’s r=0.724, CI: 0.647-0.786, P<0.001). At the outset of this investigation, Calgary’s northeast (NE) was recognized to have disproportionate COVID-19 case burden (Figure 5A-C). Accordingly, three specific representative NE neighborhoods that were assessed to have COVID-19 incidence risk of very high (NE1), high (NE2) and medium (NE3) were prioritized for monitoring. From December 2020 to January 2021 (second wave), mean SARS-CoV-2 RNA concentrations were higher at NE1 compared to NE2 and NE3 (mean values were 1340, 625 and 287 copies/ml, respectively; P <0.001, Kruskal-Wallis test) (Figure 3B). During this period, multiple comparative analyses between these sites showed NE1 to differ significantly from NE2 and NE3 (adjusted P value = 0.017 and <0.001, respectively; Dunn’s multiple comparisons test). During the trough following the second wave (i.e., from February to March 2021), new clinical cases and wastewater signals were lower for all three neighborhoods (Figure 5A-C). Beginning in March 2021 wastewater testing began for two additional neighborhoods in the southeast quadrant, where socioeconomic data indicated higher household incomes (Figure 5D and 5E; Figure 6). Significant differences in the SARS-CoV-2 RNA concentration between NE and SE neighborhoods were observed (P= 0.001, Kruskal-Wallis test) (Figure 3B). Specifically, the signal for the NE1 site was significantly higher than NE2, NE3 and SE2 sites (Adjusted P value = 0.034, 0.001 and 0.019, respectively, Dunn’s multiple comparisons test). No differences were observed for other pairwise comparisons between neighborhoods during that period (data not shown). In mid-April 2021 during Calgary’s third wave of COVID-19, wastewater testing of an additional neighborhood in the geographically distinct SW associated with high household income was brought online (Figure 5F; Figure 6). During the third wave, the SARS-CoV-2 RNA signal for NE1 was higher than NE2, SE2 and SW1 sites (adjusted P value = 0.001, 0.018 and 0.001, respectively, Dunn’s multiple comparisons test) (Figure 3B). No differences were observed for other pairwise comparisons between neighborhoods (data not shown). In general, among all neighborhoods, the NE1 location had the highest incidence of COVID-19 cases as was reflected by the greatest abundance of SARS-CoV-2 signal detected in wastewater (Figure 5A).

**Figure 5.**
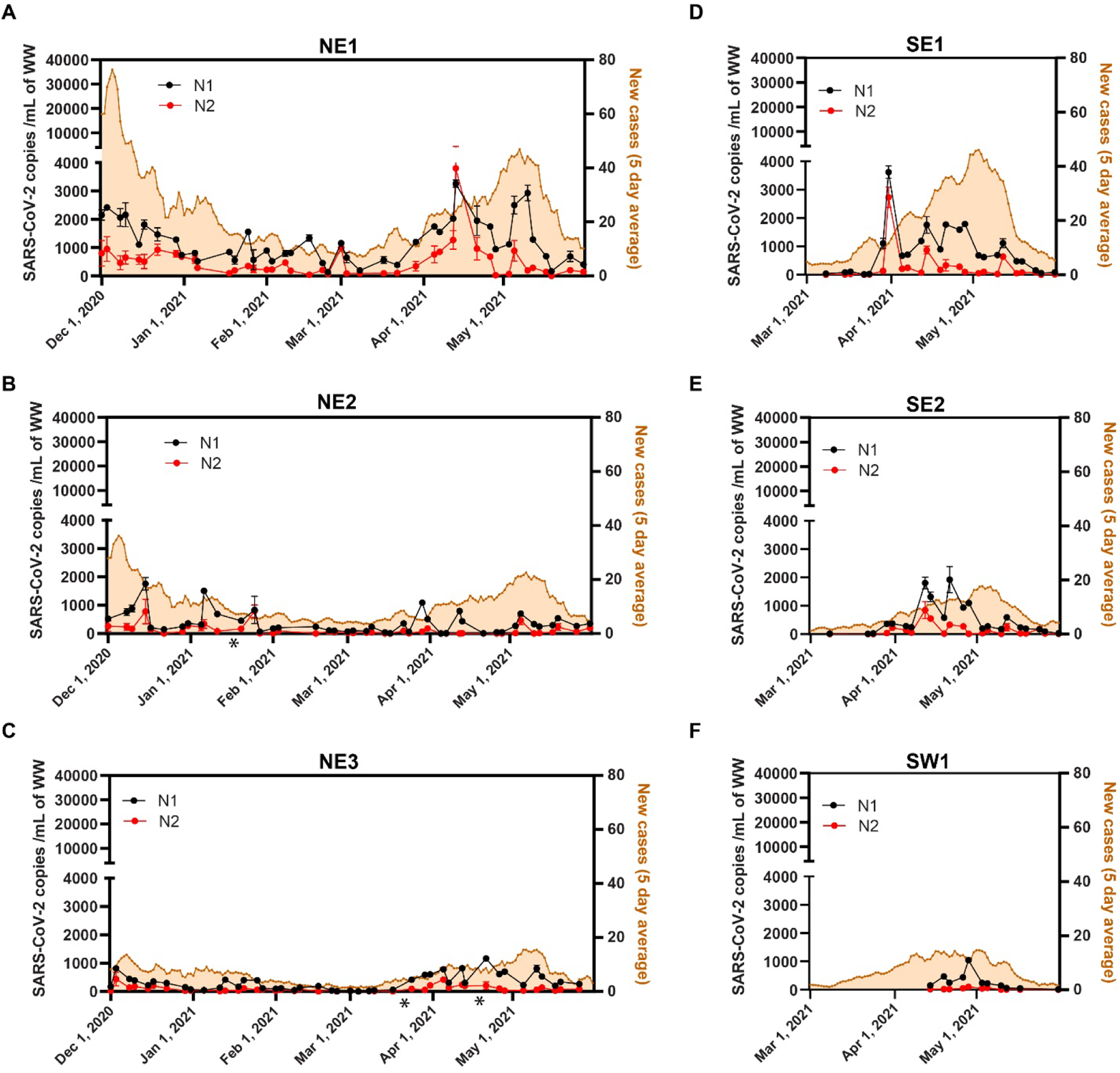
SARS-CoV-2 RNA concentration in six targeted neighborhoods in Calgary. Daily dynamics of COVID-19 new cases (5-day average) defined to the three-digit postal code level (orange line) and abundance of SARS-CoV-2 RNA signal based on N1 (black line) and N2 (red line) assays in wastewater samples. Neighborhood names correspond to the geographic quadrant of the city (northeast = NE, etc.): **A.** NE1, **B.** NE2 and **C.** NE3. South side of the city: **D.** SE1, **E.** SE2 and **F.** SW1. The number of new cases for each neighborhood were accounted by postal code data corresponding to clinically identified cases. The sampling period varied between the different neighborhoods, as shown on the x axes: NE1, NE2 and NE3 sites were sampled twice weekly from December 1, 2020 to May 31, 2021; SE1 and SE2 sites were sampled twice weekly from March 8, 2021 to May 31, 2021. SW1 was sampled twice weekly from April 14, 2021 to May 31, 2021. Error bars correspond to the standard deviations of the three technical replicates. Asterisks (*) denotes dates that were not included in the analysis (i.e., NE2 (18/01/2021), NE3 (22/03/2021) and NE3 (19/04/2021)) due to the presence of inhibitors.

**Figure 6.**
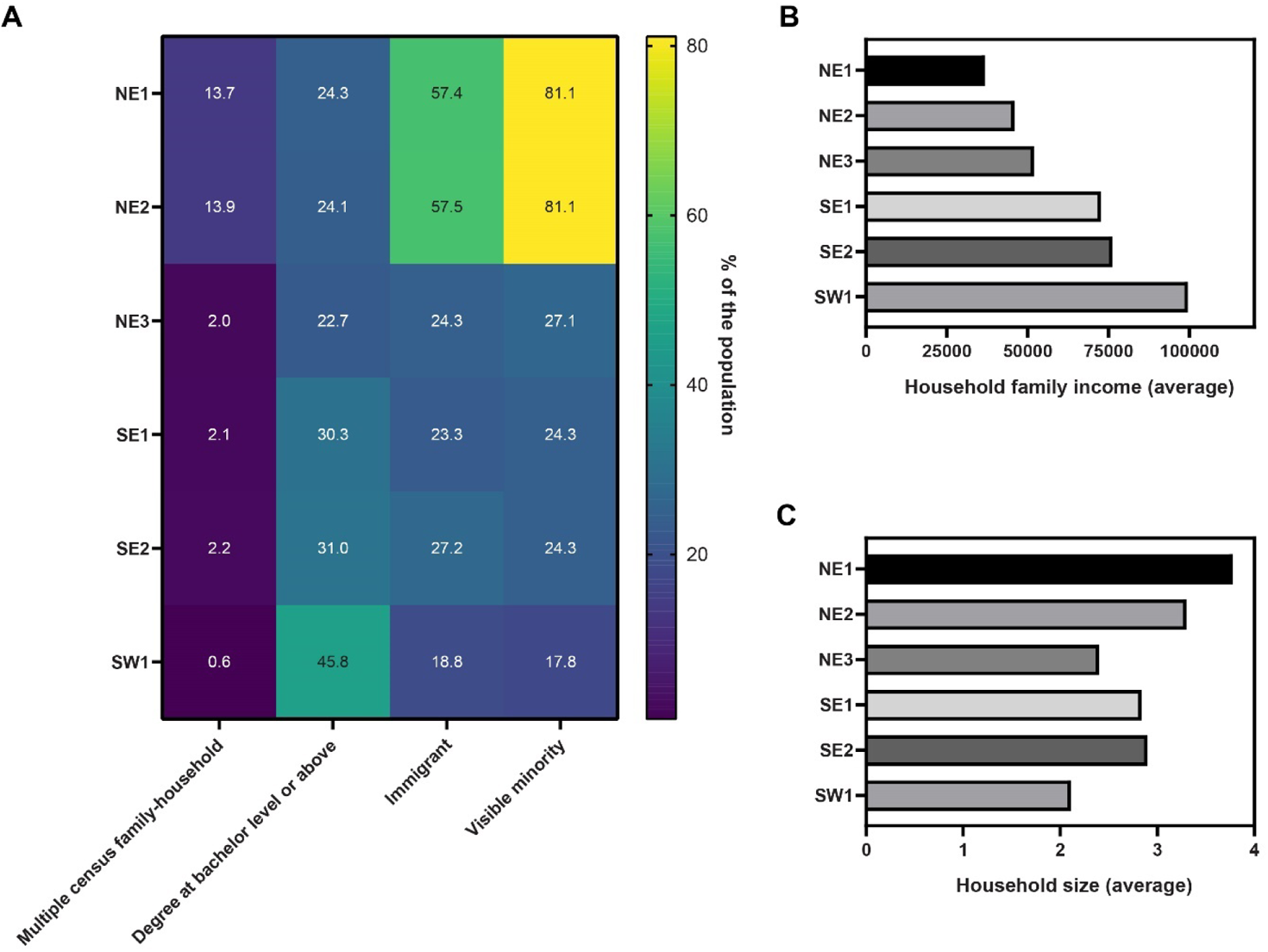
Ethno-demographic and socioeconomic status (SES) indicators for neighborhoods monitored for SARS-CoV2 in wastewater. Ethno-demographic and SES data for each neighborhood were obtained from 2016 Canadian Census data (Statistics-Canada, 2016) using postal codes that corresponded for each neighborhood-level sewershed catchment that was sampled. Family-unit (% multiple-census family households), education-level (% university certificate, diploma or degree at bachelor level or above), immigration status (% immigrant to Canada) and member of a visible minority (% visible minority), **B.** Income (measured as; average household income) and **C.** Average household size.

We observed the strongest correlations between the number of clinically identified cases (defined to the three-digit postal code level) and WW measurements at the level of the city, and WWTP relative to neighborhoods (Figure 3C and 3D). A positive correlation between the N1 signal and new cases was identified for each of the NE neighborhoods and SE2 when analyzed individually (Table 2), but not for SE1 and SW1 where fewer datapoints were collected (although the same trends evident). When SARS-CoV-2 was measured with the N2 assay, the SARS-CoV-2 signal only correlated with the number of new cases at the NE3 site (Supplemental Material, Table 4S).

To better understand the impact of nested sampling for wastewater surveillance of SARS-CoV-2, Pearson correlation analysis were conducted with all data points for all WWTP and neighborhood data. When all three WWTPs were assessed together, we observed that as new cases increased, the wastewater-signal measured with the N1 assay also increased (Pearson’s r=0.7288, CI: 0.6605-0.7851, P<0.001) (Figure 3C). Similar results were observed when N2 derived data was evaluated (Pearson’s r=0.787, CI: 0.7321-0.8331, P<0.001). When neighborhoods were assessed together (i.e., combining NE1, NE2, NE3, SE1, SE2 and SW1) the correlation was slightly lower (N1; Pearson’s r=0.6452, CI: 0.5532-0.7217, P<0.001 (Figure 3D) or N2 data; Pearson’s r= 0.3457, CI: 0.2132-0.4658, P<0.001; respectively) (Figure 3C vs D).

We sought to compare the technical success of neighborhood WBE relative to the gold-standard of WWTP (Supplementary Material, Table 3S). Failure to procure samples from neighborhoods owing to mechanical issues occurred at a higher rate than at WWTP; 26/191 (13.8%) vs 14/223 (6.3%), P=0.012. Once samples were collected, PCR-inhibition related to matrix issues were infrequent in both WWTP and neighborhoods. Only four samples out of the 414 samples subjected to molecular analysis (0.95%) were excluded due to inhibition, and this was not observed disproportionate in neighborhoods relative to WWTP (3/191 vs 1/223) (Fisher’s exact test, P= 0.339).

## 4. Discussion

WBE is an efficient and unbiased population health monitoring tool capable of recognizing changing incidence of COVID-19 at a population level, regardless of symptoms or the availability of clinical testing. Fundamental to this technology is its inclusivity and comprehensive nature. WBE surveillance combined with real-time data sharing is an independent and transparent tool that complements government reported clinical testing to understand the COVID-19 pandemic and can inform public health decision making (Kondilis et al., 2021; Ronquillo et al., 2020; The Lancet Digital, 2020).

Other groups worldwide have reported on their own local experience using this emerging technology at the level of WWTP (D’Aoust et al., 2021; Peccia et al., 2020; Wannigama et al., 2021; Wurtzer et al., 2020). These studies universally report strong correlations between wastewater SARS-CoV-2 RNA and clinical variables (including diagnosed cases, hospitalizations and COVID-19 associated deaths). The potential for WBE data to identify changes in community disease dynamics prior to clinical diagnoses is consistently reported, with the lead time varying from study to study. It directed from WWTPs has intrinsic advantages, including ease of access, existing technical expertise, and infrastructure. However, as WWTP include large and diverse geographic catchments, they do not provide information or insight on the complex geospatial distribution of COVID-19 in a community. Since the start of the pandemic, neighborhoods with racialized communities and of lower socioeconomic status have consistently been impacted disproportionally (Patel et al., 2020). Accordingly, many have advocated for the early adoption of nodal sewershed sampling to provide more granular (and potentially actionable) data for public health and education interventions, resource distribution and vaccine-targeting (Barrios et al., 2021; McClary-Gutierrez et al., 2021). From a technical perspective however, the resources required for neighborhood nodal monitoring are considerable, including the need for accurate maps of the sewer network, computer modelling of sewershed drainage areas, sufficiently granular flow measurements, and sufficiently detailed census and public health data for the corresponding resident population. Neighborhood-level WBE programs must also accommodate the complex and dynamic realities related to the feasibility and safety of accessing the desired field sampling points. Municipal sewershed networks are not designed for this kind of sampling, and in some instances the best-situated sample access points from an epidemiological perspective may be in locations (e.g., busy roads) that are relative inaccessible for high-frequency sampling events.

With our nodal based monitoring of Calgary, we observed the nucleocapsid N1-region to be more sensitive than N-2, at the level of the WWTP, individual neighborhoods and the composite city-wide metric. Other groups have reported similar trends in that the N1-target is the most sensitive marker in WWTP studies (Acosta et al., 2021; Hong et al., 2021; Huang et al., 2021; Medema et al., 2020), although this has not been universal (Ai et al., 2021).

Relative to WWTP sampling, sub-catchment scale WBE (i.e., neighborhoods) is in its infancy. This study uniquely correlates zonal SARS-CoV-2 RNA data with the collective demographics of residents in corresponding neighborhoods as determined through detailed census data. Neighborhoods with the highest SARS-CoV-2 signals in wastewater were areas where the greatest number of COVID-19 diagnosed were reported clinically. Our study took place at a time in which vaccine availability was extremely limited in Calgary’s general population (% fully vaccinated March 31^st^ – 2.5%, April 30^th^ – 7%, and May 31^st^ – 9.5%) (Alberta, 2021). As individuals with a learned immune response (either through vaccination or natural infection) are more likely to have mild symptoms that are less likely to prompt clinical testing (Harder et al., 2021; Tang et al., 2021) – in the future monitoring for breakthrough COVID-19 infections through nodal WBE could prove more important as the pandemic wanes to identify persistent pockets of ongoing, unidentified transmission.

Uniquely, we were able to correlate cases with SES and ethnodemographic indicators using comprehensive 2016 census data from Statistics Canada (Statistics-Canada, 2016) detailed to the postal code level (Figure 6). For example, neighborhoods with the highest SARS-CoV-2 in wastewater were associated with lower household income (Figure 6B) and higher numbers of residents per household (Figure 6C).Real-time monitoring with transparent reporting of neighborhood wastewater SARS-CoV-2 will provide an additional tool to help engage marginalized communities and ensure that the threat of COVID-19 is related to the unique circumstances of each community (Ahmed et al., 2021; Quinn and Andrasik, 2021; Salma and Giri, 2021).

It is not surprising that we observed mechanical complications that prevented neighborhood sample collection to occur at a slightly higher frequency than at WWTPs – although samples were still successfully collected 86% of the time. The dynamic nature of remote fieldwork including the requirement of raising and lowering autosamplers into the sewershed through manholes, creates the opportunities for increased wear and resultant equipment failure relative to the more controlled environment and infrastructure of a static WWTP monitoring program. We did not find that samples collected at the neighborhood level were more likely to be subjected to matrix problems or inhibition, with the <1% of samples excluded from analysis being independent of sampling location. In fact, we had expected matrix effects to be more prominent proximal in the sewershed as dispersion and dilution are more likely to occur at the level of WWTP.

Others too have begun exploring sub-catchment monitoring. Targeted surveillance from five manholes through Lincoln, Nebraska USA was used to infer ZIP code level regional variation in SARS-CoV-2 activity (Barrios et al., 2021). Rubio-Acero *et al*., longitudinally monitored six areas in Munich, Germany - capturing 32% of the total city’s population, demonstrating congruence between wastewater SARS-CoV-2 levels and regionally identified cases and regional distribution of specific variants of concern (VOC) (Rubio-Acero et al., 2021). Similarly, a study from Nice France, assessed wastewater between October and March 2021 and used qPCR quantification and targeted-PCR tiling with ARCTIC network primers and Oxford Nanopore Sequencing Devices to understand regional dynamics in COVID-19 (Rios et al., 2021). The authors observed that in areas purported to have residents of lower SES, wastewater SARS-CoV-2 RNA values were frequently higher than those of higher SES and specific VOC were concentrated in distinct geographic areas (Rios et al., 2021). Mota et al., monitored fifteen sewershed sub-catchments in addition to two central WWTP to perform nodal sampling in Belo Horizonte, Brazil to comprehensively capture portions of the community (in an area where clinical testing was limited) in order to track the effectiveness of public health measures (Mota et al., 2021). This revealed that the highest levels of SARS-CoV-2 were present in areas with the most vulnerable populations (i.e., favelas or shanty towns). Yaniv et al. (2021), used neighborhood level sampling to monitor the city of Ashkelon, Israel at the end of their first wave, during a period of low viral transmission to document viral shedding even in the absence of clinically diagnosed cases. They reported stronger signals at the level of sub-catchments relative to WWTP that they attributed to potential dilution effects which occurred downstream at the WWTP relative to individual neighborhoods. In contrast, we report the opposite – WWTP level data correlated with case data (limited to three-digit postal codes) to a greater degree than neighborhood-level data in Calgary - possibly owing to the wastewater sewershed’s separation from the stormwater system.

Within the confines of 3-digit postal code case-correlations, our data demonstrates the strongest correlations between cases and SARS-CoV-2 signal considered across the entire city > WWTP > individual neighborhoods. Significant correlations at the neighborhood scale were observed in NE1, NE2 and NE3 that were monitored for the longest time frame and where clinical cases were higher than in the other three neighborhoods. There are limitations to assigning clinical case residence based on three-digit postal codes, as these boundaries do not necessarily align precisely with sewershed drainage networks. Sampling at larger WWTP and city-wide scales result in these potential ‘overlap artifacts’ fading away, resulting in stronger correlations for these larger catchments and populations. This limitation can in principle be accounted for in future efforts where granular WBE data is sought. Novel strategies that maintain individuals’ anonymity while facilitating precise attribution of diagnosed cases to addresses can be developed with the guidance of local ethics boards (Hrudey et al., 2021). Other factors may also contribute to improved correlation of SARS-CoV-2 wastewater signals with clinically identified cases at larger geographies. In areas of lower SES and higher occupancy per residence (i.e., crowding), secondary infection rates within a household are higher potentially concentrating the wastewater signal (Koureas et al., 2021; Madewell et al., 2021). This is consistent with evidence that the adherence to physical and social distancing recommendations are correlated with SES indicators (Jay et al., 2020).

A major limitation of wastewater surveillance is that the signal recorded is only contributed to by individuals who defecate in a particular sewershed catchment. Indeed, most studies either show no or extremely low levels of SARS-CoV-2 excreted in urine (with ≥10 more RNA shed/g in stool relative to urine; (Jones et al., 2020)). Toileting pattens are therefore a critical determinant when ascribing wastewater data to populations. While academic studies on toileting are scarce (Heaton et al., 1992), a multitude of non-academic surveys exist which highlight a potential limitation in trying to understand patterns at increasingly granular scales. Most individuals report once-daily defecation patterns, with the majority occurring in the morning at home (although considerable variation exists). This is the rationale by which wastewater data is ascribed to a diagnosed cases home residence for COVID-19 WBE. Obviously individuals still defecate outside the home, most commonly at their place of employment, but almost half of employees report a level of discomfort defecating at work (Wong et al., 2019). Further, the usage of public restroom facilities for defecation is more likely if respondents could be assured of a single occupancy restroom (Healthline, 2017). Further refinements to COVID-19 WBE studies may need to link clinical cases not only with sewersheds of primary residences but also with primary sites of employment to capture toileting variables more comprehensively.

Understanding the natural history of SARS-CoV-2 RNA excreted into wastewater is also necessary when correlating measured signals with clinical cases. Confounding the already complicated wastewater matrix is the fact that there is likely large human differences in amount, duration and time course of fecal SARS-CoV-2 RNA shedding, potentially influenced by age, immune status, and further affected by viral factors including VOC fitness (Frampton et al., 2021; Medema et al., 2020; Mousazadeh et al., 2021). Despite the prolonged fecal shedding reported by human studies (mean 21 days), wastewater studies generally most strongly correlate

with new daily cases, with a 4- to 7-day lead time (Nemudryi et al., 2020; Peccia et al., 2020). Monitoring the wastewater of three tertiary care hospitals in Calgary using the same methods as described here, it was observed that large increases in SARS-CoV-2 RNA were attributable to even single or small clusters of hospital acquired transmission despite the presence of dozens of hospitalized patients on-site, further suggesting that shedding is greatest at symptom onset (Acosta et al., 2021).

There are several other limitations in this work that warrant discussion. Most notably, we were limited in the frequency and number of samples that were analyzed per week per location, primarily related to the cost associated with collection from neighborhoods. More frequent sample collection would be useful for early detection of outbreaks or rapid changes in the trajectory of the SARS-CoV-2 RNA signal in wastewater. For more granular WBE to be successful, new strategies independent of the dangerous and confined spaces hazards of municipally installed autosamplers in local sewersheds need to be realized. Our study focuses primarily on detection and trend determination for SARS-CoV-2 RNA copies in wastewater samples obtained at different catchment scales; future research should examine aspects such as sensitivity and lead time estimation.

## 5. Conclusions

Wastewater-based monitoring of SARS-CoV-2 RNA across a range of geospatial scales correlated well with clinically diagnosed COVID-19 cases in Calgary. Composite flow-adjusted signals from three individual WWTPs enabled a single city-wide metric that correlated strongly with clinically diagnosed cases geographically constrained to the three-digit postal code level. The more granular neighborhood-level sub-catchment data showed a significant correlation, but the limitations of three-digit postal code case attribution reduced sensitivity. We believe that the actionability of WBE ultimately will be greatest when performed at granular levels. However, at present nodal-WBE requires a more refined understanding / modeling of the system dynamics, because the system is “noisier” (more heterogeneous data) at smaller monitoring scales.

## Author contributions

**Nicole Acosta:** Formal analysis, Investigation, Methodology, Validation, Data Curation, Writing - Original Draft, Writing - Review & Editing. **María A. Bautista**: Methodology, Validation, Investigation. **Barbara J. Waddell**; Methodology, validation, investigation, data curation. **Janine McCalder**. Methodology, Validation, investigation. **Alexander Buchner Beaudet**; Investigation. **Lawrence Man**; Investigation. **Puja Pradhan**; Investigation. **Navid Sedaghat**; Investigation. **Chloe Papparis**; Investigation. **Andra Bacanu**; Investigation. **Jordan Hollman**; Methodology, Validation. **Alexander Krusina**; Data Curation, Visualization. **Danielle Southern**; Data Curation, Visualization, **Tyler Williamson**; Data Curation, Visualization. **Carmen Li**; Methodology. **Srijak Bhatnagar**; Formal analysis, visualization. **Sean Murphy**; Investigation. **Jianwei Chen**; Methodology. **Darina Kuzma**; Validation, Methodology. **Jon Meddings**; Project administration, funding acquisition. **Jia Hu**; Funding acquisition. **Jason L. Cabaj**; Funding acquisition. **John M. Conly;** Formal analysis, Writing - Review & Editing. **Norma J. Ruecker;** Project administration, Funding acquisition, Investigation. **Gopal Achari:** Conceptualization, Formal analysis, Writing - Review & Editing. **M. Cathryn Ryan:** Conceptualization, Formal analysis, Writing - Review & Editing. **Kevin Frankowski:** Conceptualization, Formal analysis, Writing - Review & Editing. **Casey RJ Hubert:** Conceptualization, Formal analysis, Funding acquisition, Writing - Review & Editing. **Michael D. Parkins:** Conceptualization, Formal analysis, Funding acquisition, Supervision, Writing - Review & Editing.

## Data Availability

Data is accessible from reasonable requests to the authors.

Real-time data on SARS-CoV-2 RNA levels has been updated on the Cumming School of Medicine Centre for Health Informatics Website. https://covid-tracker.chi-csm.ca

## Acknowledgments

The investigators are appreciative of the staff at Alberta Health Services DIMR (Analytics, Data Integration, Measurement & Reporting) for data collection and sharing. We are particularly grateful to the City of Calgary and in particular members of Water Quality Services Infrastructure Planning and Wastewater Treatment for their excellent support on provision of samples, data, and infrastructure information. The authors gratefully acknowledge assistance from Dr Rhonda Clark for program administration and management. The graphical abstract was created with BioRender.

## Abbreviations

BCoV: Bovine coronavirus
Cq: quantification cycle
NTC: no-template control
PMMoV: Pepper mild mottle virus
SARS-CoV-2: Severe Acute Respiratory Syndrome Coronavirus 2
SES: Socioeconomic status
VOC: variants of concern
WBE: wastewater-based epidemiology
WWTP: wastewater treatment plant

