## Supplemental Files for "Longitudinal SARS-CoV-2 RNA Wastewater Monitoring Across a Range of Scales Correlates with Total and Regional COVID-19 Burden in a Well-Defined Urban Population"

**1.** **SUPPLEMENTARY METHODS**

**1.1 Wastewater sampling**

ISCO portable samplers were programmed to collect and store 24-hour composite wastewater samples as follows: At WWTP-1, 100 ml aliquots were collected for each 3200 m^3^ of flow – ensuring equivalence to WWTP-2 and 3. At WWTP-2, 70 ml aliquots were collected every 15 minutes. At WWTP-3, 150 ml aliquots were collected every 30 minutes. In neighborhoods, 100 ml aliquots were collected every 15 minutes.

**1.2. Quality assurance and quality control**

All samples were preserved on ice and were processed within one day of arrival at the Advancing Canadian Wastewater Assets laboratory. To preserve the SARS-CoV-2 RNA signal in the wastewater, each wastewater sample was subsampled upon receipt and mixed with NaCl and EDTA to final concentrations of 4 M and 1 mM, respectively. Each wastewater sample was spiked with BCoV as surrogate external control (Bovilis® Coronavirus Vaccine; Merck, Catalogue #151921) to a final concentration of 2500 TCID50/ml before performing the 4S protocol (Whitney et al., 2021). BCoV spike-in control monitoring enabled the detection of inhibitors in samples and ensured the efficiency of nucleic acid extraction.

**1.3. RT-qPCR analysis**

N1 and N2 RT-qPCR reaction mixtures had a final volume of 20 μl and contained 1 × TaqMan™ Fast Virus 1-Step Master Mix (Applied Biosystems), final concentrations of 500 nM for both forward and reverse primers (i.e., N1 or N2, (CDC, 2020)), a final concentration of 125 nM for TaqMan probes (CDC, 2020), and 5 µl RNA template. Quantification of BCoV was performed as described previously (Acosta et al., 2021) using final concentrations of 200 nM for forward and reverse primers and 125 nM of the TaqMan probe (Decaro et al., 2008). Thermal cycling conditions for different assays are summarized in Tables S1 and S2.

A human fecal control marker, pepper mild mottle virus (PMMoV) replicase gene (Kitajima et al., 2018), was used as a second control (internal) to confirm potential for inhibition when BCoV values were low. Samples where BCoV and PMMoV were markedly reduced were flagged and investigated for potential inhibition. To determine the presence of RT-qPCR inhibitors within a sample, we followed a spike and dilution methodology. Inhibition was evaluated by processing the samples using the described Sewage, Salt, Silica, and SARS-CoV-2 (4S) protocol without the addition of the internal control. Then, nucleic acids extracts were spiked by a known amount of BCoV RNA (7.6 x 107 copies/ml) or UltraPure™ DNase/RNase-Free Distilled Water. Mixes of BCoV RNA with nucleic acids extracts or distilled water were diluted in 1:5 and 1:10 dilutions and were used as template for the BCoV RT-qPCR assay. Samples were deemed inhibited if the Cq value was greater than two cycles from the expected Cq value for the amplification of the uninhibited control (i.e., UltraPure™ DNase/RNase-Free Distilled Water (Invitrogen)) (Staley et al., 2012). Samples that were affected by PCR inhibition were excluded from further analyses.

No-template controls (NTCs; UltraPure™ DNase/RNase-Free Distilled Water (Invitrogen)) and extraction blank controls were included in triplicate in every RT-qPCR assay for all targets (i.e., N1, N2, BCoV and PMMoV). Similarly, a positive control for the N1 and N2 assays (i.e., 2019-nCoV_N positive control (Integrated DNA Technologies, Catalogue # 10006625) and for the BCoV-PMMoV assays (i.e., pMCSG53 vector plasmid containing a 482 bp in length of a region of the transmembrane (M) gene of the BCoV and a region of the replicase gene of the PMMoV (Acosta et al., 2021)) were included in triplicate. To determine the limit of detection of the N1 and N2 RT-qPCR assays, two-fold serial dilutions of the TWIST AR-S SARS-Cov-2 RNA control 2 (Twist Bioscience, MN 908947.3; 8 to 0.0625 genomic copies/ µl) was performed and analyzed in octuplicate and the LOD was determined as the last dilution where the relative repeatability standard deviation (RSDr) of the replicates was ≤ 33% (Del Gaudio et al., 2012).

**1.4. Demographic and data collection**

WWTP-1 is the largest WWTP within Calgary, treating 80% of the total daily wastewater flow. WWTP-2 and WWTP-3 collectively treat wastewater from the south catchment area and as such, the number of attributable COVID-19 cases were the same for both for data analysis purposes. Characteristics of each community under WBE monitoring were established through linkage to 2016 Canadian Census data (Pinault et al., 2020; Statistics-Canada, 2016). Socioeconomic status (SES) and ethno-demographic indicators under investigation included: Income (measured as average household income), Education Level (% university certificate, diploma or degree at bachelor level or above), Family Unit (average household size; % multiple-census family households refers to whether in a single household there is more than one census family), Immigration Status (% immigrant to Canada: includes persons who are, or who have ever been, landed immigrants or permanent residents) and status as a member of a Visible Minority (% persons, other than Aboriginal peoples, who are non-Caucasian in race or non-white in colour).

**2.** **SUPPLEMENTARY RESULTS**

**2.1. Assay performance and controls**

A total of four samples were excluded secondary to the presence of qPCR inhibitors, one WWTP sample (February 15^th^, 2021, from WWTP-1) and three neighborhood samples (January 1^st^ from NE2; March 22^nd^ and April 19^th^ from NE3). SARS-CoV-2 RNA signal was detected in 98% of the wastewater samples analyzed. As previously reported (Acosta et al., 2021), N1 demonstrated greater sensitivity relative to N2 for detection and quantification of SARS-CoV-2 in wastewater. Of the 410 samples tested using N1- and N2-assays, 375 samples were positive for both targets, and 402 were positive only for N1. No samples were positive for N2 and negative for N1. The mean RT-qPCR efficiency for the N1 and N2 assay was 116.6% and 100.2% respectively. Standard curves used to quantify SARS-CoV-2 RNA had a mean slope and mean R^2^ value of -3.02 and 0.69 for the N1 assay and -3.34 and 0.94 for the N2 assay. This resulted in limits of detection being 0.27 and 0.61 genomic copies per reaction for N1 and N2, respectively. BCoV spike-in control estimation varied between 1.1 x 10^5^ and 6.7 x 10^6^ copies/ml without significant differences between different WWTP and neighbourhood sampling sites (P=0.6648, Kruskal-Wallis test). To test whether inter-sample differences in BCoV load could affect the observed measurements of N1 and N2, Pearson correlations between N1 or N2 and BCoV signals were performed. Neither N1 or N2 RNA signal correlated with BCoV concentrations (P > 0.05). NTCs and extraction blanks controls were negative for all targets (i.e., N1, N2 and BCoV) in all assays that were performed.

**2.2 Correlating wastewater data with public health action**

During our longitudinal temporal monitoring program for SARS-CoV-2 in wastewater (starting at the end of Calgary’s “first wave” in May 2020) a series of public health mandates were implemented and rescinded. At the study’s initiation in the trough of wave 1, the number of newly diagnosed clinical cases was stable and remained low throughout the summer, with a median of 2.3 cases per 100,000 people from late June to mid-September 2020. Similar trends were observed in flow-corrected wastewater data (Figure 4A). In the fall, levels of SARS-CoV-2 in wastewater began to increase from October to mid-December 2020, coinciding with an increase of clinically diagnosed new COVID-19 cases in Calgary. To deal with this “second wave”, increasingly stringent restrictions were implemented on November 24 and December 8 (Figure 4A, see blue-shaded periods). Thereafter, sharp decreases in both the SARS-CoV-2 wastewater signal and clinically diagnosed cases were observed (Figure 4A, first purple-shaded area). In response to this drop in COVID-19, on February 8, 2021, some of the second wave restrictions were partially lifted, with further relaxations being implemented on March 8, 2021 (Figure 4A). These events were associated with a “third wave” of increases in wastewater SARS-CoV-2 signals and subsequently clinical cases, leading to another introduction of restrictions on April 9, 2021. To mitigate the major surge of new cases during this third wave (Figure 4A, gray-shaded area), additional new restrictions were introduced on May 7, 2021. Following this, there were reductions in the SARS-CoV-2 signal (Figure 4A, second purple-shaded area) and the number of new COVID-19 cases.

**SUPPLEMENTARY TABLES**

**Table 1S.** Thermal cycling conditions for each target gene used in the study.

| Target gene | N1 or N2 | | | | BCoV and PMMoV | | |
| --- | --- | --- | --- | --- | --- | --- | --- |
|  | **Temperature** | **Time** | **Cycles** | **Temperature** | | **Time** | **Cycles** |
| UNG incubation | - | - | - | 25 ℃ | | 2 min | 1 |
| Reverse transcription | 50 ℃ | 5 min | 1 | 53 ℃ | | 10 min | 1 |
| RT inactivation /initial denaturation | 95 ℃ | 20 sec | 1 | 95 ℃ | | 2 min | 1 |
| Denature | 95 ℃ | 3 sec | 45 | 95 ℃ | | 3 sec | 40 |
| Anneal / extend | 55 ℃ | 30 sec | 45 | 60 ℃ | | 30 sec | 40 |

**Table 2S.** Primers and probes for each target used in the study.

| Target | Name | Oligonucleotide Sequence (5’ – 3’) | Reference |
| --- | --- | --- | --- |
| N1 | 2019-nCoV_N1-F | GACCCCAAAATCAGCGAAAT | (CDC, 2020) |
|  | 2019-nCoV_N1-P | ACCCCGCATTACGTTTGGTGGACC |  |
|  | 2019-nCoV_N1-R | TCTGGTTACTGCCAGTTGAATCTG |  |
| N2 | 2019-nCoV_N2-F | TTACAAACATTGGCCGCAAA | (CDC, 2020) |
|  | 2019-nCoV_N2-P | ACAATTTGCCCCCAGCGCTTCAG |  |
|  | 2019-nCoV_N2-R | GCGCGACATTCCGAAGAA |  |
| BCoV | BCoV-F | CTGGAAGTTGGTGGAGTT | (Decaro et al., 2008) |
|  | BCoV-Pb | CCTTCATATCTATACACATCAAGTTGTT |  |
|  | BCoV-R | ATTATCGGCCTAACATACATC |  |
| PMMoV | PMMV-FP1-rev | GAGTGGTTTGACCTTAACGTTTGA | (Zhang et al., 2005) |
|  | PMMV-Probe1 | CCTACCGAAGCAAATG | (Haramoto et al., 2013) |
|  | PMMV-RP1 | TTGTCGGTTGCAATGCAAGT |  |

**Table 3S.** Weekly sample collection from WWTPs and neighborhoods.

| Dates | Sampling/ week^β^ | WWTP* | | | Neighbourhood* | | | | | |
| --- | --- | --- | --- | --- | --- | --- | --- | --- | --- | --- |
|  |  | **WWTP-1** | **WWTP-2** | **WWTP-3** | **NE1** | **NE2** | **NE3** | **SE1** | **SE2** | **SW1** |
| Jun 29 to Jul 5, 2020 | 3 | 1 | 1 | 1 |  |  |  |  |  |  |
| Jul 6 to Jul 12, 2020 | 3 | 1 | 1 | 1 |  |  |  |  |  |  |
| Jul 13 to Jul 19, 2020 | 3 | 0 | 0 | 0 |  |  |  |  |  |  |
| Jul 20 to Jul 26, 2020 | 3 | 1 | 1 | 1 |  |  |  |  |  |  |
| Jul 27 to Aug 2, 2020 | 3 | 1 | 1 | 1 |  |  |  |  |  |  |
| Aug 3 to Aug 9, 2020 | 3 | 0 | 0 | 1 |  |  |  |  |  |  |
| Aug 10 to Aug 16, 2020 | 3 | 1 | 1 | 0 |  |  |  |  |  |  |
| Aug 17 to Aug 23, 2020 | 3 | 1 | 1 | 1 |  |  |  |  |  |  |
| Aug 24 to Aug 30, 2020 | 3 | 0 | 0 | 0 |  |  |  |  |  |  |
| Aug 31 to Sep 6, 2020 | 3 | 1 | 1 | 1 |  |  |  |  |  |  |
| Sep 7 to Sep 13, 2020 | 3 | 1 | 1 | 1 |  |  |  |  |  |  |
| Sep 14 to Sep 20, 2020 | 3 | 0 | 0 | 0 |  |  |  |  |  |  |
| Sep 21 to Sep 27, 2020 | 3 | 1 | 1 | 1 |  |  |  |  |  |  |
| Sep 28 to Oct 4, 2020 | 9 | 1 | 3 | 3 |  |  |  |  |  |  |
| Oct 5 to Oct 11, 2020 | 9 | 3 | 3 | 3 |  |  |  |  |  |  |
| Oct 12 to Oct 18, 2020 | 9 | 3 | 3 | 3 |  |  |  |  |  |  |
| Oct 19 to Oct 25, 2020 | 9 | 3 | 3 | 3 |  |  |  |  |  |  |
| Oct 26 to Nov 1, 2020 | 9 | 3 | 3 | 3 |  |  |  |  |  |  |
| Nov 2 to Nov 8, 2020 | 3 | 1 | 1 | 1 |  |  |  |  |  |  |
| Nov 9 to Nov 15, 2020 | 3 | 1 | 1 | 1 |  |  |  |  |  |  |
| Nov 16 to Nov 22, 2020 | 3 | 1 | 1 | 1 |  |  |  |  |  |  |
| Nov 23 to Nov 29, 2020 | 3 | 1 | 1 | 1 |  |  |  |  |  |  |
| Nov 30 to Dec 6, 2020 | 3 / 6 | 1 | 1 | 1 | 2 | 1 | 2 |  |  |  |
| Dec 7 to Dec 13, 2020 | 3 / 6 | 1 | 1 | 1 | 2 | 2 | 2 |  |  |  |
| Dec 14 to Dec 20, 2020 | 3 / 6 | 1 | 1 | 1 | 2 | 2 | 2 |  |  |  |
| Dec 21 to Dec 27, 2020 | 3 / 6 | 1 | 1 | 1 | 1 | 1 | 1 |  |  |  |
| Dec 28 to Dec 31, 2020 | 3 / 6 | 1 | 1 | 1 | 2 | 2 | 2 |  |  |  |
| Jan 1 to Jan 3, 2021 | 0 | 0 | 0 | 0 | 0 | 0 | 0 |  |  |  |
| Jan 4 to Jan 10, 2021 | 3 / 6 | 1 | 1 | 1 | 2 | 2 | 1 |  |  |  |
| Jan 11 to Jan 17, 2021 | 6 / 6 | 2 | 2 | 2 | 0 | 1 | 2 |  |  |  |
| Jan 18 to Jan 24, 2021 | 6 / 6 | 2 | 2 | 2 | 2 | 1 | 2 |  |  |  |
| Jan 25 to Jan 31, 2021 | 6 / 6 | 2 | 2 | 2 | 2 | 2 | 2 |  |  |  |
| Feb 1 to Feb 7, 2021 | 6 / 6 | 2 | 2 | 2 | 2 | 2 | 2 |  |  |  |
| Feb 8 to Feb 14, 2021 | 6 / 6 | 2 | 2 | 2 | 2 | 0 | 2 |  |  |  |
| Feb 15 to Feb 21, 2021 | 6 / 6 | 1 | 2 | 2 | 1 | 1 | 1 |  |  |  |
| Feb 22 to Feb 28, 2021 | 6 / 6 | 2 | 2 | 2 | 2 | 2 | 2 |  |  |  |
| Mar 1 to Mar 7, 2021 | 6 / 6 | 2 | 2 | 2 | 2 | 2 | 2 |  |  |  |
| Mar 8 to Mar 14, 2021 | 6 / 10 | 2 | 2 | 2 | 1 | 2 | 2 | 1 | 1 |  |
| Mar 15 to Mar 21, 2021 | 6 / 10 | 2 | 2 | 2 | 1 | 2 | 1 | 2 | 0 |  |
| Mar 22 to Mar 28, 2021 | 6 / 10 | 2 | 2 | 2 | 1 | 2 | 1 | 2 | 2 |  |
| Mar 29 to Apr 4, 2021 | 6 / 10 | 2 | 2 | 2 | 1 | 2 | 2 | 2 | 2 |  |
| Apr 5 to Apr 11, 2021 | 6 / 10 | 2 | 2 | 2 | 2 | 2 | 2 | 2 | 2 |  |
| Apr 12 to Apr 18, 2021 | 6 / 11 | 2 | 2 | 2 | 2 | 2 | 2 | 2 | 2 | 1 |
| Apr 19 to Apr 25, 2021 | 6 / 12 | 2 | 2 | 2 | 1 | 1 | 1 | 2 | 2 | 2 |
| Apr 26 to May 2, 2021 | 6 / 12 | 2 | 2 | 2 | 2 | 2 | 2 | 2 | 2 | 2 |
| May 3 to May 9, 2021 | 6 / 12 | 2 | 2 | 2 | 2 | 2 | 1 | 2 | 2 | 2 |
| May 10 to May 16, 2021 | 6 / 12 | 2 | 2 | 2 | 2 | 2 | 2 | 2 | 2 | 2 |
| May 17 to May 23, 2021 | 6 / 12 | 2 | 2 | 2 | 2 | 2 | 2 | 2 | 2 | 1 |
| May 24 to May 30, 2021 | 6 / 12 | 2 | 2 | 2 | 1 | 1 | 1 | 2 | 2 | 0 |
| May 31 to Jun 6, 2021 | 3 / 6 | 1 | 1 | 1 | 1 | 1 | 0 | 1 | 1 | 1 |
| Total | | **72** | **75** | **75** | **43** | **44** | **44** | **24** | **22** | **11** |

^β^ Planned number of sampling events. *When samples were not collected at an individual sampling site during each week of monitoring the explanation is provided using a colour legend. Orange: no sample was collected for mechanical failures; Purple: no sample collected due to civic holidays; Gray: sample collected successfully but excluded owing to low signal in internal controls.

**Table 4S.** Correlations between daily COVID-19 cases and SARS-CoV-2 gene copies/ml in different sampling locations using the N2 assay

| Location | SARS-CoV-2 gene copies/ml vs new cases (rolling 5-day average)* | | |
| --- | --- | --- | --- |
|  | **r** | **P value** | **95 % CI** |
| WWTP-1 | 0.777 | <0.0001 | 0.664 to 0.855 |
| WWTP-2 | 0.711 | <0.0001 | 0.577 to 0.808 |
| WWTP-3 | 0.678 | <0.0001 | 0.533 to 0.785 |
| NE1 | 0.210 | 0.18 | -0.097 to 0.480 |
| NE2 | 0.233 | 0.13 | -0.069 to 0.495 |
| NE3 | 0.401 | 0.007 | 0.119 to 0.624 |
| SE1 | -0.003 | 0.987 | -0.406 to 0.4006 |
| SE2 | 0.210 | 0.348 | -0.232 to 0.580 |
| SW1 | 0.462 | 0.15 | -0.191 to 0.832 |

* New cases defined to the three-digit postal code level.
